# Reproducibility in the UK Biobank of Genome-Wide Significant Signals Discovered in Earlier Genome-wide Association Studies

**DOI:** 10.1101/2020.06.24.20139576

**Authors:** Jack W. O’Sullivan, John P. A. Ioannidis

## Abstract

With the establishment of large biobanks, discovery of single nucleotide polymorphism (SNPs) that are associated with various phenotypes has been accelerated. An open question is whether SNPs identified with genome-wide significance in earlier genome-wide association studies (GWAS) are replicated also in later GWAS conducted in biobanks. To address this question, the authors examined a publicly available GWAS database and identified two, independent GWAS on the same phenotype (an earlier, “discovery” GWAS and a later, replication GWAS done in the UK biobank). The analysis evaluated 136,318,924 SNPs (of which 6,289 had reached p<5e-8 in the discovery GWAS) from 4,397,962 participants across nine phenotypes. The overall replication rate was 85.0% and it was lower for binary than for quantitative phenotypes (58.1% versus 94.8% respectively). There was a18.0% decrease in SNP effect size for binary phenotypes, but a 12.0% increase for quantitative phenotypes. Using the discovery SNP effect size, phenotype trait (binary or quantitative), and discovery p-value, we built and validated a model that predicted SNP replication with area under the Receiver Operator Curve = 0.90. While non-replication may often reflect lack of power rather than genuine false-positive findings, these results provide insights about which discovered associations are likely to be seen again across subsequent GWAS.

## Introduction

Genome-wide association studies (GWAS) have already a long track record and have resulted in the discovery of tens of thousands of genetic associations for various traits and phenotypes. Polygenic risk scores (1), innovative drug discovery (2), and gene-editing (3) have all been enhanced, or even based on, GWAS results. Genome-wide association studies investigate the association of individual single nucleotide polymorphisms (SNPs) on a phenotype of interest (for example coronary artery diseases) (4). Most GWAS identify SNPs with, individually, small effects (4). This supports the notion that most diseases are polygenic, rather than monogenic, in nature (5).

To observe the small effect of individual SNPs, GWAS have relied on increasingly larger sample sizes (4). Recent advances have seen rapidly increasing sample sizes, particularly with the establishment of large biobanks. The most widely used and analyzed biobank in human genetics is the UK Biobank (UKBB) (6). Analyses done in the UK biobank and other similar biobanks have the opportunity not only to identify new associations but also to replicate previously proposed associations that arose from other GWAS investigations. It is not unexpected that some SNPs that were considered to be associated with a phenotype in an earlier GWAS may not be replicated in a subsequent GWAS. Even if they are replicated, their effect size may change, e.g. because of the winner’s curse phenomenon (7), where early discoveries see attenuation of their effect size when they are replicated in subsequent studies. This has implications for all scientific progress, and even patient care, (i.e. polygenic risk scores) reliant on GWAS results, if these scores include variants that have null effects or effects that are smaller than those anticipated based on their earlier discovery profile.

Although several studies have looked at SNP replication for specific phenotypes, it remains broadly unclear across phenotypes how often SNPs replicate, how this varies between binary and quantitative traits, at different p-values, across varying effect sizes, and how effect sizes change between earlier, smaller GWAS and later, larger GWAS examining the same phenotype. A most interesting comparison would be to contrast earlier GWAS versus the UK biobank, which has become a standard, widely used resource. We set out to address these questions, and, from our results, built a model to predict SNP replication.

## Methods

### Data acquisition

To determine the reproducibility of SNPs between an earlier GWAS and the UK biobank, we identified two, independent GWAS on the same trait, one without data from the UK biobank and the second being done on UK biobank data. To do this, we systematically searched a publically available database of genome-wide association studies (GWAS) (available at: https://atlas.ctglab.nl/) (8) for GWAS that had been conducted for the same trait (e.g. systolic blood pressure) first using data independent of the UK Biobank (UKBB) and then a second, independent GWAS using exclusively UKBB data. Thus a trait was eligible if there were two independent GWAS available for it; one not using UKBB data (hereafter referred to as: discovery GWAS) and one using UKBB data (hereafter: replication GWAS). All discovery GWAS occured before the replication GWAS. Further inclusion criteria was GWAS conducted in European subjects (or results available for exclusively Europeans) and GWAS with more than 50 genome-wide significant SNPs, so as to allow having a meaningful number of discoveries to be assessed for replication. More information on the GWAS database we searched and its accompanying paper (8) are available in the appendix.

### Determination of reproducibility

To determine the reproducibility of SNPs in the discovery and replication GWAS we performed three broad steps: 1. Determined overlap of SNPs between discovery and replication GWAS (via rsID) and only included SNPs shared between two GWAS cohorts. We then identified the SNPs that reached genome-wide significance (defined consistently as P<5e-8, regardless of the threshold that the original authors might have used) in the discovery GWAS - these were the SNPs we determined the reproducibility of. 2. Aligned the effect allele between the discovery and replication GWAS, and consequently inverted the effect size if effect alleles did not originally match and 3. Classified SNPs as replicated if they reached genome-wide significant (P<5e-8) in both discovery and replication GWAS and had congruent effect directions in both GWAS (e.g. odds ratio (OR) above 1 in both GWAS). All SNP effect sizes were converted to OR before reproducibility was determined via the Chinn formula (9). Thus, SNP effect sizes that were originally produced from linear models for quantitative (continuous) traits were converted to OR. Further details appear in the appendix.

### Calculating reproducibility

We calculated the replication rate for each included trait individually, for all traits collectively, and for binary (e.g. coronary artery disease) and quantitative (e.g. diastolic blood pressure) traits separately. To calculate replication rate for each individual trait we calculated a simple proportion (e.g. [number of SNPs replicated] / [number of SNPs shared between discovery and replication GWAS]). To calculate the replication rate for all traits collectively we constructed a inverse-variance meta-analysis (10) using fixed-effects. Further, we constructed similar inverse-variance meta-analysis (10) to determine the replication rate for binary and quantitative traits; including only traits recorded in a binary fashion (yes/no) or on a continuous scale, respectively. To explore the replication rate across P-values and odd ratios, we also performed meta-analysis assessing the replication of SNPs with certain P-value and OR characteristics (from the discovery GWAS). We calculated the reproducibility of SNPs across the following discovery GWAS P-value categories: 5e-8 to 5e-9, 5e-9 to 5e-10, 5e-10 to 5e-11, and <5e-11. We calculated the reproducibility of SNPs across the following discovery GWAS OR categories: 1-1.05, 1.05-1.1, 1.1-1.15, 1.15-1.2, 1.2-1.3, 1.3-1.4, >1.4.

### Quantifying the change in effect size between GWAS

To determine if a change in SNP effect size occured between the earlier, discovery GWAS and the later, replication GWAS in the UKBB we constructed a single variate linear model, with the discovery OR as the predictor variable and replication OR as the outcome variable. As stated above (see ‘*Determination of reproducibility’)*, we converted all SNP effect sizes to an OR via the Chinn formula (9). Then, to help interpret the output from this model, we converted all OR values to above 1 (using the formula 1/OR if the original SNP OR was <1) Finally we combined SNPs across all traits for the model. From the regression model, we determined the regression coefficient for the discovery OR and interpreted this coefficient as the change in OR between GWAS (e.g. a regression coefficient of 0.80 would imply that 20% decrease in OR between discovery and replication GWAS). We only quantified the change in effect size of SNPs that were replicated, and also for all SNPs that had reached genome-wide significance in the discovery GWAS, regardless of whether they were replicated or not in the replication GWAS. We performed similar analyses for binary and quantitative traits individually.

### Prediction model for SNP replication

First we constructed a multivariate logistic regression model to examine the association of our predictors (odds ratio, p-value, p-value category (as above), and trait characteristic (binary vs. quantitative) on replication. We initially split our data into test and train sets (split, randomly, by half). Using the train set, we constructed logistic regression model using the following predictors: odds ratio (numeric, not category), p-value category, and trait characteristic (binary vs. quantitative). We then tested the constructed model on the test set. We report the model’s predictive accuracy via the following metrics: sensitivity, specificity, and area under the curve (AUC) all with 95% confidence intervals. We further assessed model fit via McFadden’s R^2^.

## Results

We analysed 136,318,924 SNPs from 4,397,962 participants across nine different phenotypes (from 18 GWAS, 9 pairs) (table 1). The traits included were: asthma, systolic blood pressure (SBP), eczema, body mass index (BMI), waist circumference, hip circumference, coronary artery disease (CAD), resting pulse rate, and diastolic blood pressure (DBP). Of the 136,318,924 included SNPs, 6,289 reached genome-wide significance (P < 5e-8) in the discovery GWAS (table 1 and eTable1).

**Table 1.**

| Disease | Total sample size | Number of Genome-wide significant SNPs | Number of SNPs that are replicated (%) |
| --- | --- | --- | --- |
| Asthma | 225,309 | 889 | 494 (56%) |
| SBP | 430,797 | 110 | 107 (97%) |
| Eczema | 330,142 | 640 | 337 (53%) |
| BMI | 613,900 | 1835 | 1756 (96%) |
| Waist Circumference | 618,033 | 937 | 827 (89%) |
| Hip circumference | 598,925 | 1083 | 1043 (96%) |
| Coronary Artery Disease/IHD | 387,786 | 159 | 149 (94%) |
| Resting Heart rate/Pulse Rate | 447,198 | 549 | 547 (99%) |
| DBP | 430,806 | 87 | 83 (95%) |

Total sample size is the sample size of the discovery and replication GWAS collectively.

### Replication rate

Of the 6,289 SNPs that were genome-wide significant in the discovery cohort, 5,343 were replicated in the replication cohort (85.0%, 95% Confidence Interval (CI): 84.1% to 85.8%) (eFigure 1). Results varied substantially between binary and quantitative traits; the replication rate for exclusively binary phenotypes was 58.1% (95%CI: 55.7% to 60.4%) (eFigure 2), compared with 94.8% (95%CI: 94.2% to 95.4%) for quantitative traits (eFigure 3). The replication rate varied across the included phenotypes from 52.7% to 99.6% (Figure 1).

**Figure 1.**
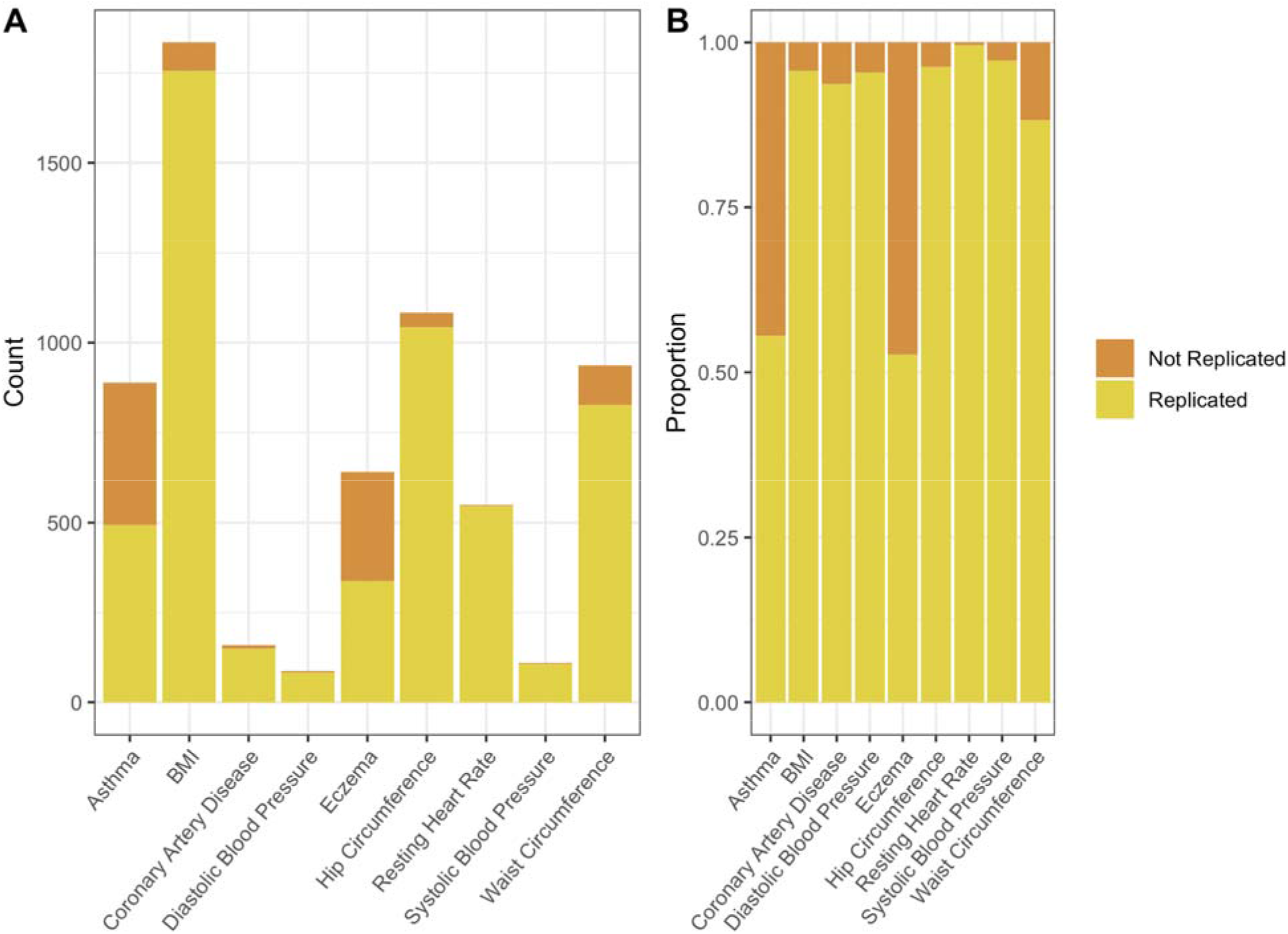

Furthermore, the replication rate varied across discovery GWAS P-values and OR (Figure 2, Figure 3, eFigure 4 and eFigure 5). As is expected, the replication rate increased as the discovery GWAS SNP P-value decreased (table 2); the highest replication was observed with a P-value <5e-11 (94% (95%CI: 93% to 95%). A less consistent pattern was observed with discovery GWAS OR, almost all OR >/= 1.2 were replicated (table 2), however a similarly large number of SNPs with a discovery OR of >1 to <1.05 were replicated (94.3% (95%CI: 93.5% to 95.0%)). This is likely due to the fact that all SNPs >1 to <1.05 were for quantitative traits, with no SNPs corresponding to binary traits (figure 4).

**Figure 2.**
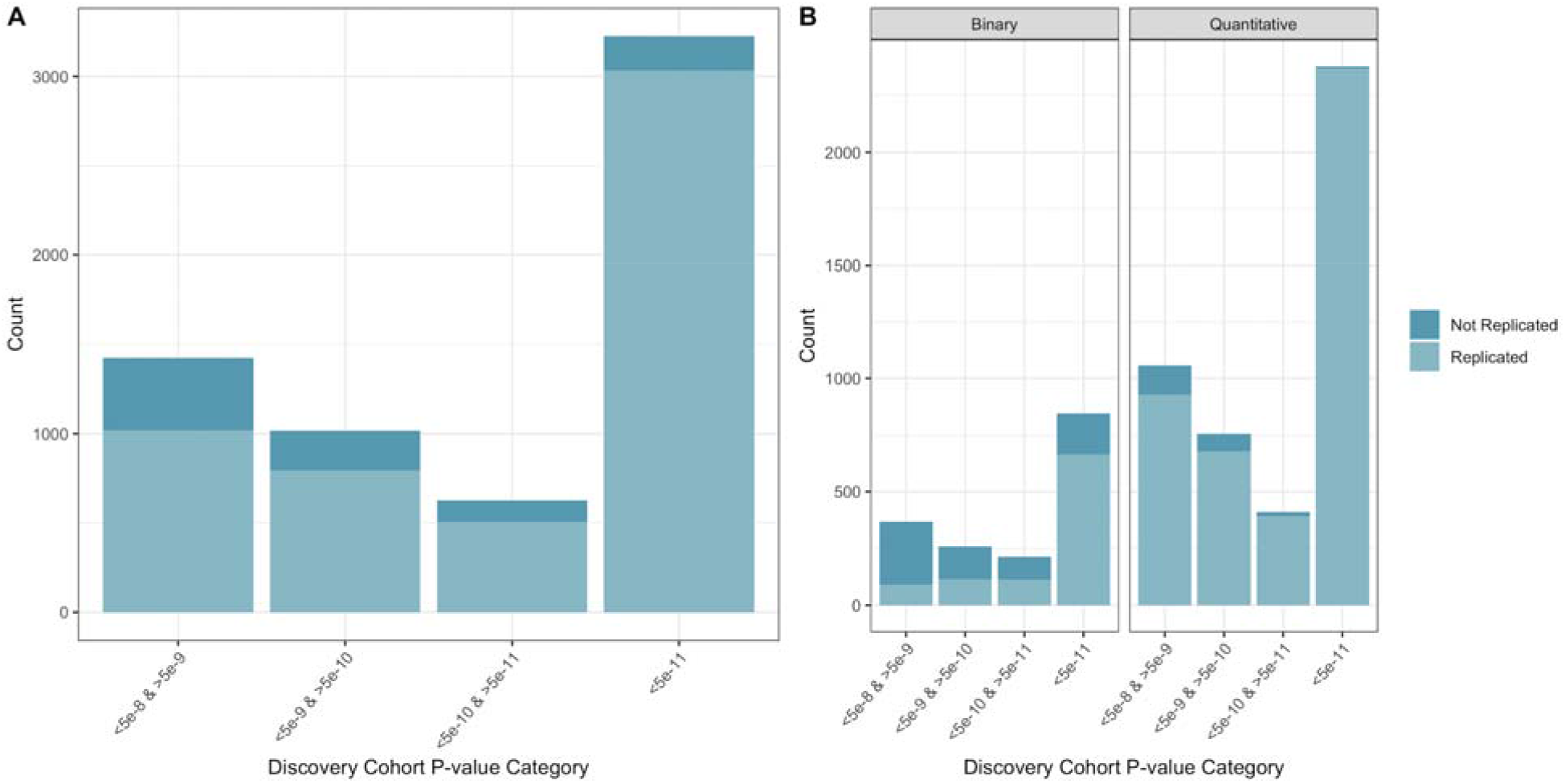

**Figure 3.**
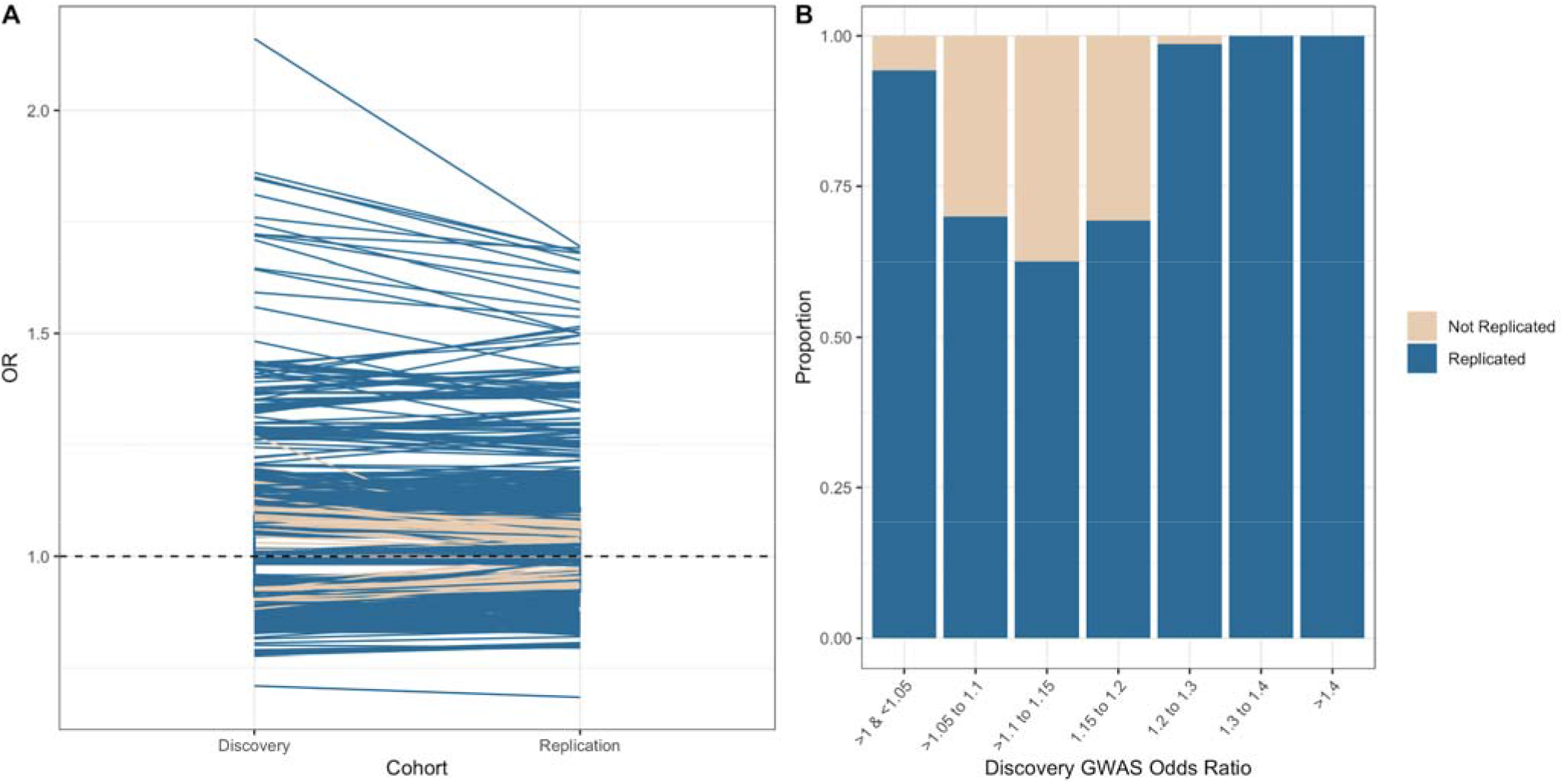

**Table 2.**

| Metric | Category | Replication rate (95%CI) |
| --- | --- | --- |
| P-Value | 5e-8 to >5e-9 | 72% (69% to 74%) |
|  | 5e-9 to >5e-10 | 78% (75% to 80%) |
|  | 5e-10 to >5e-11 | 81% (77% to 83%) |
|  | <5e-11 | 94% (93% to 95%) |
| Odds Ratio | 1-1.05 | 94.3% (93.5% to 95.0%) |
|  | 1.05-1.1 | 70.0% (66.8% to 72.9%) |
|  | 1.1-1.15 | 62.5% (59.4% to 65.6%) |
|  | 1.15-1.2 | 69.3% (64.3% to 73.9%) |
|  | 1.2-1.3 | 98.7% (91.0% to 99.8%) |
|  | 1.3-1.4 | 100%* |
|  | >1.4 | 100%* |

\* Paucity of data prevented formal meta-analysis

**Figure 4.**
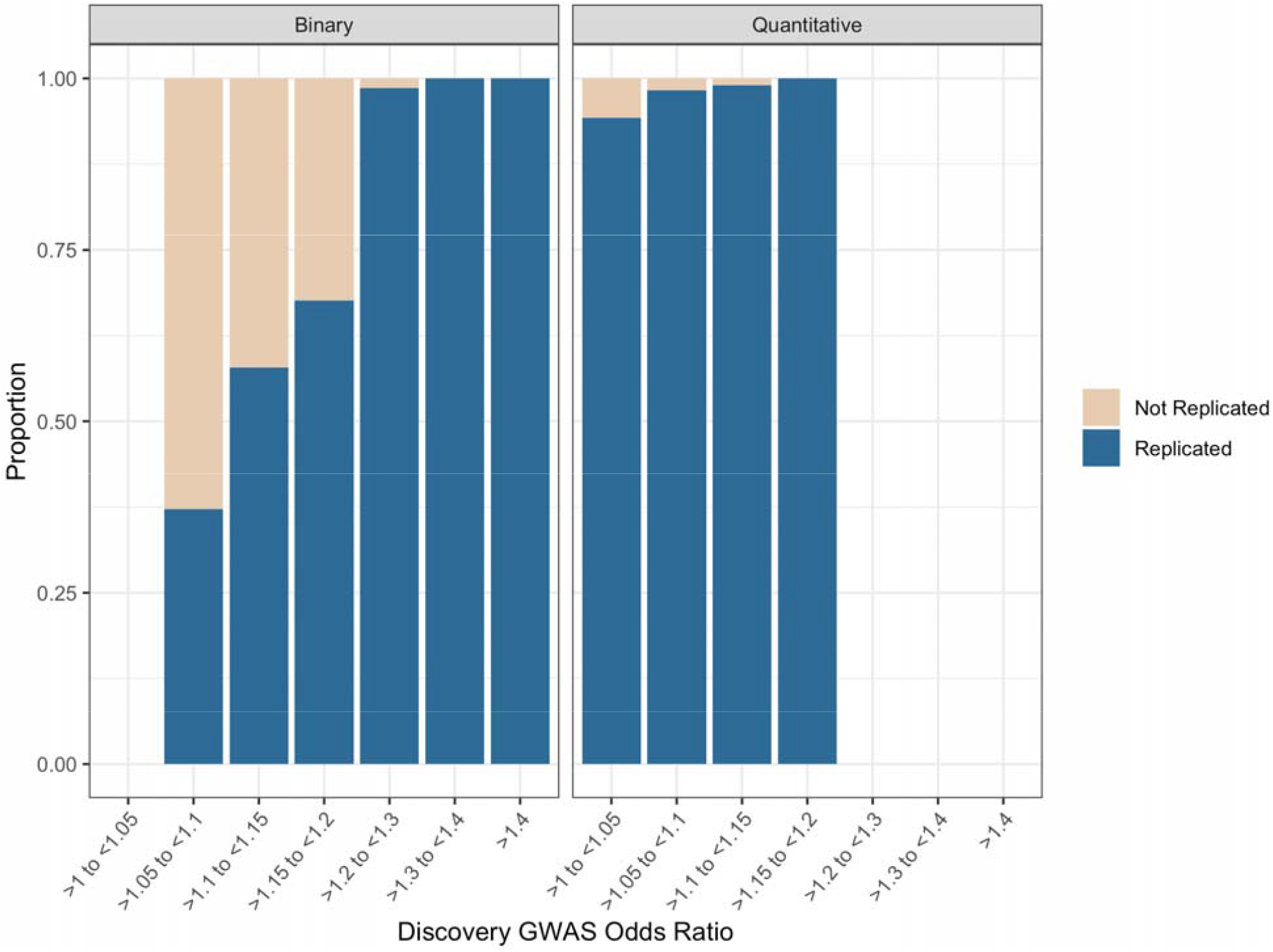

### Change in effect size between GWAS

When considering SNPs that were replicated in both cohorts, we found a 9.6% (95%CI: 8.9% to 10.2%) decrease in replicated SNP OR between discovery and replication cohorts (Figure 3), for all phenotypes collectively. This decrease in effect size was larger for binary traits (18.0% (95%CI: 16.0% to 20.0%), eFigure 6), however for quantitative traits an increase in effect size was observed (12.0% (95%CI: 11.0% to 13.0%), eFigure 6). The change in effect size varied substantially across phenotypes (eFigure 7).

When considering SNPs that reached genome-wide significance in the discovery cohort (and weren’t necessarily replicated), we found a 16.4% (95%CI: 82.8% to 84.4%) decrease in SNP OR between discovery and replication cohorts, for all phenotypes collectively. For binary traits this decrease was 13.6% (95%CI: 11.4% to 15.9%), whereas we observed a 10.9% (95%CI: 9.9% to 11.9%) increase for quantitative traits.

### Predicting SNP replication

First, from our training model the following predictors were significantly associated with SNP replication: discovery cohort SNP odds ratio (figure 4), discovery cohort trait (binary or quantitative), discovery cohort SNP p-value <5e-10 &>5e-11, and discovery cohort SNP p-value <5e-11 (both categorical variables with p-value <5e-8 &>5e-9 as reference) (eTable2). P-value as a continuous variable and p-value <5e-9 &>5e-10 were not significant (eTable2).

When we applied our training model to our test data set, we found an area under the Receiver Operator Curve (ROC) of 0.90 (95%CI: 0.89 to 0.91) corresponding to a sensitivity and specificity of 70.9% (95%CI: 69.2% to 84.5%) and 93.6% (95%CI: 80.0% to 95.6%) respectively (eFigure 8). We found a McFadden’s R^2^ of 0.33.

## Discussion

We analysed 136,318,924 SNPs from 4,397,962 participants across nine different phenotypes (18 GWAS). Of these 136,318,924 SNPs, 6,289 SNPs reached genome-wide significance in the respective discovery GWAS, of which 5,343 were replicated in their replication GWAS (85.0%, 95% Confidence Interval (CI): 84.1% to 85.8%). Replication rate varied substantially between binary and quantitative phenotypes and it was much lower in the former. Further, replication rate varied across P-value and OR of discovery GWAS SNP. We also found that SNP odds ratios (OR) decreased between discovery and replication GWAS for binary phenotypes, but increased for quantitative phenotypes. Lastly, we developed and then validated a model to predict SNP replication, and found it to be accurate (0.90 (95%CI: 0.89 to 0.91)).

### Implications

Our results have implications for the potential validity and utility of GWAS results. First, the SNP replication rate for quantitative phenotypes is very high; implying that quantitative GWAS in the UKBB had likely reached sufficient power to accurately detect all SNPs that were truly associated with a phenotype and that had been discovered by earlier GWAS efforts. The high replication rate observed for quantitative traits may also reflect the precision and relative ease in which quantitative traits can be measured. The converse of this, the likely measurement error and ultimate definition heterogeneity of binary phenotypes, may be one explanation for the relatively low rate of replication in binary phenotypes. For instance, binary phenotypes often represent complex clinical diseases that can have a) broad diagnostic criteria (e.g. angina, and myocardial infarction are often captured under “Coronary Artery Disease”) and b) are defined via an array of data sources, of varying quality. The UKBB, for instance, defines their phenotypes with ICD codes based on linked electronic health records (EHR).(6) While this probably represents the best current method to define phenotypes in large cohorts, EHR data is “messy” and likely to include some “administrative and clinical error” (11). An improvement in the phenotyping in data used for GWAS of binary phenotypes is likely to result in improved SNP replication. This may be even more crucial for phenotypes where we saw low replication rates, e.g. eczema.

While the quality of phenotyping will eventually improve, in the meantime the modest replication rate we observed poses questions about the best way to utilize current binary phenotype GWAS results. On the one hand, it is encouraging that much scientific progress has been accomplished with current binary GWAS. For instance, polygenic risk scores based on current binary GWAS have been shown to accurately predict complex, common phenotypes (1,12,13). With improved phenotyping, it seems plausible that these scores may continue to improve. Nevertheless, in the meantime there may be other ways to enhance current binary GWAS results for polygenic risk scores. First, our results clearly show a superior replication rate with quantitative phenotypes. These quantitative phenotypes are often more in line with physiological processes (e.g. systolic blood pressure) than clinical diseases (e.g. coronary artery disease). As such, future GWAS that directly use metabolomic data as outcomes (such as protein expression) are likely to, similarly, have higher accuracy than clinical disease phenotypes. Future research merging metabolomic outcomes and GWAS may be a useful addition to our scientific knowledge. Second, almost all SNPs for binary traits with an OR >/= 1.2 were replicated, whereas the majority of SNPs with an OR below 1.2 were not replicated and this may reflect lack of power in the replication dataset. Of note, many of the replication UKBB datasets that we considered here did not use the full UKBB data, and power is likely to improve as complete biobank data are used and many biobanks are combined.

### Limitations in comparison to previous literature

We were surprised to find only nine phenotypes where two GWAS had been conducted in truly independent participants and where inclusion or not of UKBB data was a distinguishing feature. It is plausible that further independent GWAS on the same traits exist, although this seems unlikely given the thorough and systematic search we performed of the GWAS atlas (8). It is, however, likely that more GWAS are available, but they contain overlapping samples between GWAS (i.e. two GWAS of the same phenotype are not truly independent as they contain similar cohorts of participants), aren’t of sufficient quality to be included in the GWAS Atlas, are conducted in a non-European population, or have not made their summary statistics available. A earlier study (14) reports building a model for SNP replication using GWAS for over 50 phenotypes, although it is unclear what, if any, measures were taken to determine if these numerous GWAS were truly independent i.e. did not include overlapping participants. Also, this study validated their model in two, small GWAS of one trait. Furthermore, this study didn’t actually quantify a SNP replication rate, nor did they stratify their results by binary and quantitative phenotypes. A further limitation of our study is that we didn’t include other SNP features, ideally we would have liked to include, for instance, minor allele frequency as a predictor in our model. However, this data was sparsely available in the replication (non-UKBB) GWAS. Lastly, it should be acknowledged that large disease-specific consortiums generally qualitatively describe the replication of SNPs as their consortium increases. Our study quantifies this formally and, importantly, quantifies replication across more than one phenotype.

### Future research

We have identified a number of future research priorities. First, improving the phenotyping of binary phenotypes seems to be a priority for GWAS. Second, to facilitate an assessment of SNP replication, future independent cohorts are likely required. Many efforts to do this are already underway (e.g. AllofUs cohort and Millions Veteran Program).

## Conclusions

The replication of SNPs discovered from GWAS was high for quantitative phenotypes. Genome-wide Association Studies appear to be entirely sufficient to detect SNPs associated with quantitative traits. For binary traits, however, the replication rate is modest. We have built a simple prediction model that can accurately ascertain SNP replication in later GWAS. It may be of use for researchers and clinicians that utilize GWAS results.

## Data Availability

All data is publicly available at: https://atlas.ctglab.nl/

## Notes

### Competing Interest Statement

The authors have declared no competing interest.

### Funding Statement

The lead author (JOS) was supported by an NIH T32 grant, otherwise, there is no specific funding.

### Author Declarations

All data is publicly available as summary statistics from UK Biobank

## References

1. O’Sullivan JW, Shcherbina A, Justesen JM, et al. Combining clinical and polygenic risk improves stroke prediction among individuals with atrial fibrillation. MedRxiv. 2020;(https://www.medrxiv.org/content/10.1101/2020.06.17.20134163v1.article-info)

2. Shu L, Blencowe M, Yang X. Translating GWAS Findings to Novel Therapeutic Targets for Coronary Artery Disease. Front Cardiovasc Med. 2018;5:56.

3. Wu S, Zhang M, Yang X, et al. Genome-wide association studies and CRISPR/Cas9-mediated gene editing identify regulatory variants influencing eyebrow thickness in humans. PLoS Genet. 2018;14(9):e1007640.

4. Tam V, Patel N, Turcotte M, et al. Benefits and limitations of genome-wide association studies. Nat. Rev. Genet. 2019;20(8):467–484.

5. Lambert SA, Abraham G, Inouye M. Towards clinical utility of polygenic risk scores. Hum. Mol. Genet. 2019;28(IR2):R133–R142.

6. Sudlow C, Gallacher J, Allen N, et al. UK biobank: an open access resource for identifying the causes of a wide range of complex diseases of middle and old age. PLoS Med. 2015;12(3):e1001779.

7. Xiao R, Boehnke M. Quantifying and correcting for the winner’s curse in genetic association studies. Genet. Epidemiol. 2009;33(5):453–462.

8. Watanabe K, Stringer S, Frei O, et al. A global overview of pleiotropy and genetic architecture in complex traits. Nat. Genet. 2019;51(9):1339–1348.

9. Chinn S. A simple method for converting an odds ratio to effect size for use in meta-analysis. Stat. Med. 2000;19(22):3127–3131.

10. Barendregt JJ, Doi SA, Lee YY, et al. Meta-analysis of prevalence. J. Epidemiol. Community Health. 2013;67(11):974–978.

11. Sudlow C. Ascertaining health outcomes through linking across the UK to NHS datasets covering a wide range of diseases. UKBB; 2017.

12. Inouye M, Abraham G, Nelson CP, et al. Genomic Risk Prediction of Coronary Artery Disease in 480,000 Adults: Implications for Primary Prevention. J. Am. Coll. Cardiol. 2018;72(16):1883–1893.

13. Abraham G, Malik R, Yonova-Doing E, et al. Genomic risk score offers predictive performance comparable to clinical risk factors for ischaemic stroke. Nat. Commun. 2019;10(1):5819.

14. Gorlov IP, Moore JH, Peng B, et al. SNP characteristics predict replication success in association studies. Hum. Genet. 2014;133(12):1477–1486.

